# Incidence and Economic Burden of Retinal Tears in the United States

**DOI:** 10.1101/2022.10.29.22281697

**Authors:** Kamil Taneja, Michael Joseph Diaz, Tanisha Taneja, Karan Patel, Sai Batchu, Solomon Oak, Alex Zhang, Aditya Joshi, Urvish K. Patel

## Abstract

**Purpose:** To characterize retinal tears (RTs) and calculate the economic burden of RTs that present to the emergency department (ED) in the US.

**Methods:** We used a large national ED database to retrospectively analyze RTs that presented to the ED from 2006 to 2019. Using extrapolation methods, national estimates of RT incidence, demographics, comorbidities, disposition, inpatient (IP) costs, and ED costs were calculated.

**Results:** During the time period between 2006 and 2019, 15,841 patients presented to the ED with RT as the primary diagnosis. RT incidence was stable at 8.2 per million US population (95% Confidence Interval (CI): 5.3 - 21.0) in this time period. Most patients were males, Caucasian, paid with private insurance, and admitted to EDs in the Northeast. The most common comorbidities were hypertension (19%), a history of cataracts (15%), and diabetes (7.2%).

During this time period, RTs costs added up to more than $79 million and $33 million in the ED and IP settings, respectively. Mean per-patient ED and IP costs increased by 145% (p=0.0008) and 86% (p=0.0047), respectively.

**Conclusion:** Despite the stable incidence of RTs, RTs place a significant economic burden to the healthcare system, which increases yearly. We recommend physicians and policy makers to work together to pass laws that could prevent the increasing healthcare costs.

## Introduction

A retinal tear (RT) occurs when the posterior vitreous tugs on the inner limiting membrane of the retina. Age^1^, gender^2^, myopia^3^, and retinitis pigmentosa^4^ are considered risk factors for posterior vitreous detachment (PVD) and RTs. When a horseshoe RT (a full-thickness RT) occurs, vitreous fluid accumulates under the retina, which can lead to a rhegmatogenous retinal detachment (RRD). Even though RTs are rare, RT treatment and recovery is a significant source of emotional and financial stress.

To our knowledge, there is no nation-wide analysis of the epidemiology of RTs. Most of these studies are retrospective chart reviews that look at treatment outcomes and occurrence of future RTs after treatment.^5–10^ 15% of all PVD patients present with a symptomatic RT and RTs occur in 70% of patients with PVD and vitreous hemorrhage.^5^ 12% of PVD-related RT patients can have future RTs, which can be asymptomatic.^8^ Approximations of RTs can be done by looking at other similar conditions such as giant RTs (GRTs), which are rare RTs that encompass more than 90 degrees of the retina, and RRDs, a common consequence of RTs. GRTs are estimated to have an incidence of 0.9-11.4 per million population^7^ and RRDs have an incidence from 91 to 170 per million population.^11^ Due to the fact that GRTs have a similar pathogenesis to RTs, it is likely that RTs are commonly idiopathic like GRTs.^7^ Overall, there is a large gap in our knowledge of the epidemiology of RTs and we can only make conclusions based on other similar conditions.

Along with the lack of understanding of the epidemiology of RTs, there is very little data on the economic burden of RTs. The rising costs of the healthcare field has been well documented.^12,13^ Briefly, the rising costs have been attributed to rising service intensity, an aging population, and an increasing population.^12^ Increases in ED charges has also been demonstrated.^12,14^ To our knowledge, there is no research on the economic burden of RTs. Although it has been shown that there has not been an increase in mean inpatient (IP) costs for eye trauma, there is very little data for specific eye trauma diagnosis.^15^ Regardless of the little research done on the economic burden of eye trauma, it is possible that ED and IP costs are increasing.^16^

In this study, we aim to fill in the gaps in our understanding of the epidemiology of RTs. We characterize the RT patient population, calculate annual RT incidence, patient outcomes, and calculate costs with a national ED database.

## Materials and Methods

### Dataset

The National Emergency Department Sample (NEDS) is a database sponsored by the Healthcare Cost and Utilization Project (HCUP). It is an all-payer database that includes data from nearly 1000 EDs in the US. For each patient, characteristics of the hospital-owned ED where the patient presented to (e.g. geographic location, trauma center designation, teaching status), patient characteristics (age, gender, diagnosis, primary payer method), and disposition (ED encounter outcome, inpatient admission status, cost) are included. This database uses the International Classification of Diseases (ICD) to describe the primary diagnosis of a patient (which is defined as the diagnosis that all care is based on) and associated diagnoses. From 2006 to the first three quarters of 2015, the ICD-9 system was used. From the last quarter of 2015 to 2019, the ICD-10 system was used. More information about the NEDS can be found in the following link:https://www.hcup-us.ahrq.gov/nedsoverview.jsp.

NEDS data was handled according to the tenets of the Declaration of Helsinki and the HCUP data use agreement. To adhere to the HCUP data use agreement, estimates less than or equal to 10 were not reported to maintain patient confidentiality.

### Design and Analysis

We conducted a retrospective longitudinal study to identify ED visits with a primary diagnosis of a retinal tear in the years 2006, 2009, 2012, 2015, 2017, and 2019. To do this, we created a database with patients that had a primary diagnosis of a retinal tear (ICD-9 Codes:361.01-361.05, 361.31-361.33 and ICD-10 Codes:H33.00-H33.059, H33.30-H33.339). National estimates were calculated by the NEDS-supplied sampling weights. Descriptive statistics were calculated for retinal tear patient demographics and hospital characteristics. Annual incidence of retinal tears was calculated by using data supplied from the US Census Bureau. Yearly cumulative and mean per-visit emergency department and inpatient costs were calculated for each year. Costs were adjusted for inflation based on the January 2021 US dollar, using the Consumer Price Index for Hospital Services from the US Bureau of Labor Statistics. Within the retinal tear patient group, we also analyzed patients that presented with a retinal tear and a RRD (ICD-9 Codes:361.01-361.05 and ICD-10 Codes:H33.00-H33.059). A p-value less than 0.05 was considered significant.

## Results

### Incidence

In the study time period, 22,128 patients presented to the ED with a RT. 15,841 of these patients had a primary diagnosis of RT. There was not a significant chance in retinal tear incidence from 2006 to 2019. The average incidence in this time period was 8.2 per million US population (range: 5.3 - 21.0 per million US population) (Figure 1A). Out of all of the primary RT patients, 10,577 (66.8%) presented with a RRD. The proportion of retinal tear patients with a RRD decreased from 81.2% to 63.3% (p=0.0017) in the study time period (Figure 1B).

**Figure 1:**
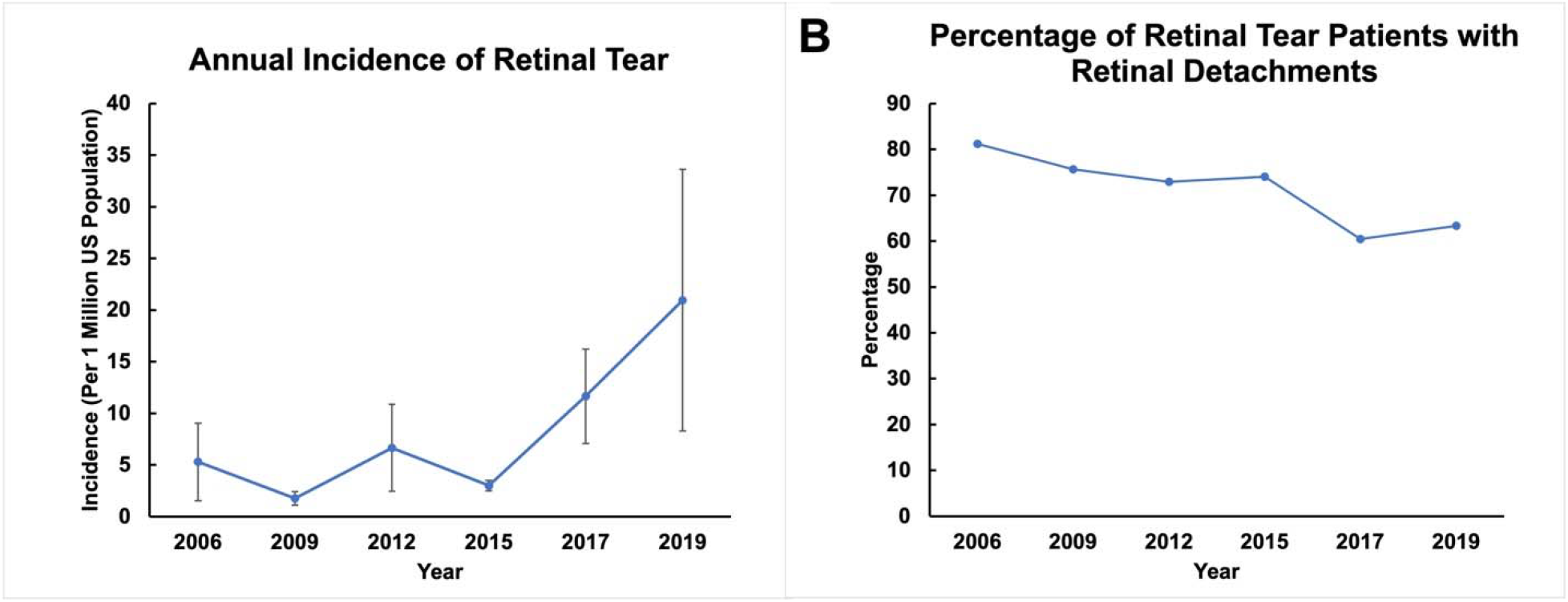
Annul trends of retinal tears. A: Annual incidence of primary retinal tears. B: Annual percentage of retinal tear patients with a rhegmatogenous retinal detachment. Error bars represent standard error.

### Demographics

The majority of RT patients were male (61.0%), White (50.3%), had private insurance (49.3%), were admitted to northeastern hospitals (53.3%), and from the highest income quartile (28.3%). The most prevalent comorbidities included hypertension (19.0%), history of cataracts (14.8%), and diabetes (7.2%). Less common comorbidities included hyperlipidemia (5.3%), obesity (2.2%), degenerative myopia (1.3%), myopia (2.5%), glaucoma (3.2%), tobacco usage (6.9%), overweight (1.4%), and alcohol usage (0.3%). 826 (5.2%) patients were admitted to the inpatient setting. A comprehensive list of patient demographics related to retinal tears can be seen in Supplementary Table 1.

### Management

Most patients treated with a primary RT were treated and discharged without IP admission (91.5%). Elderly patients (at least 65 years of age) had the highest rate of routine discharge from the ED (92.3%) and children (0-10 years of age) had the lowest rate of routine discharge (84.8%). 5.2% of patients were admitted. Middle age (45-64 years of age) had the highest rate of IP admission and elderly patients the lowest rate (3.6%) of IP admission. Discharge against medical advice (0.6%), transfers to short-term hospitals or other facilities (2.1%), and home health care (0.6%) were more rare.

### Charges and Costs

Over this time period, retinal tears lead to an inflation-adjusted cumulative ED cost of $79,476,223 and IP cost of $33,295,452. Annual total ED costs significantly increased by a factor of 10.6 from $3.7 million in 2006 to $43.1 million in 2019 (p=0.0475) (Figure 2A). Annual IP costs averaged $5.5 million per year and did not significantly change during this time period (p=0.14) (Figure 2B). Mean ED costs significantly changed by 145% from $2,569 in 2006 to $6,302 in 2019 (p=0.0008) (Figure 2C). Mean IP costs significantly changed by 86% from $23,979 in 2006 to $44,499 in 2019 (p=0.0047) (Figure 2D).

**Figure 2:**
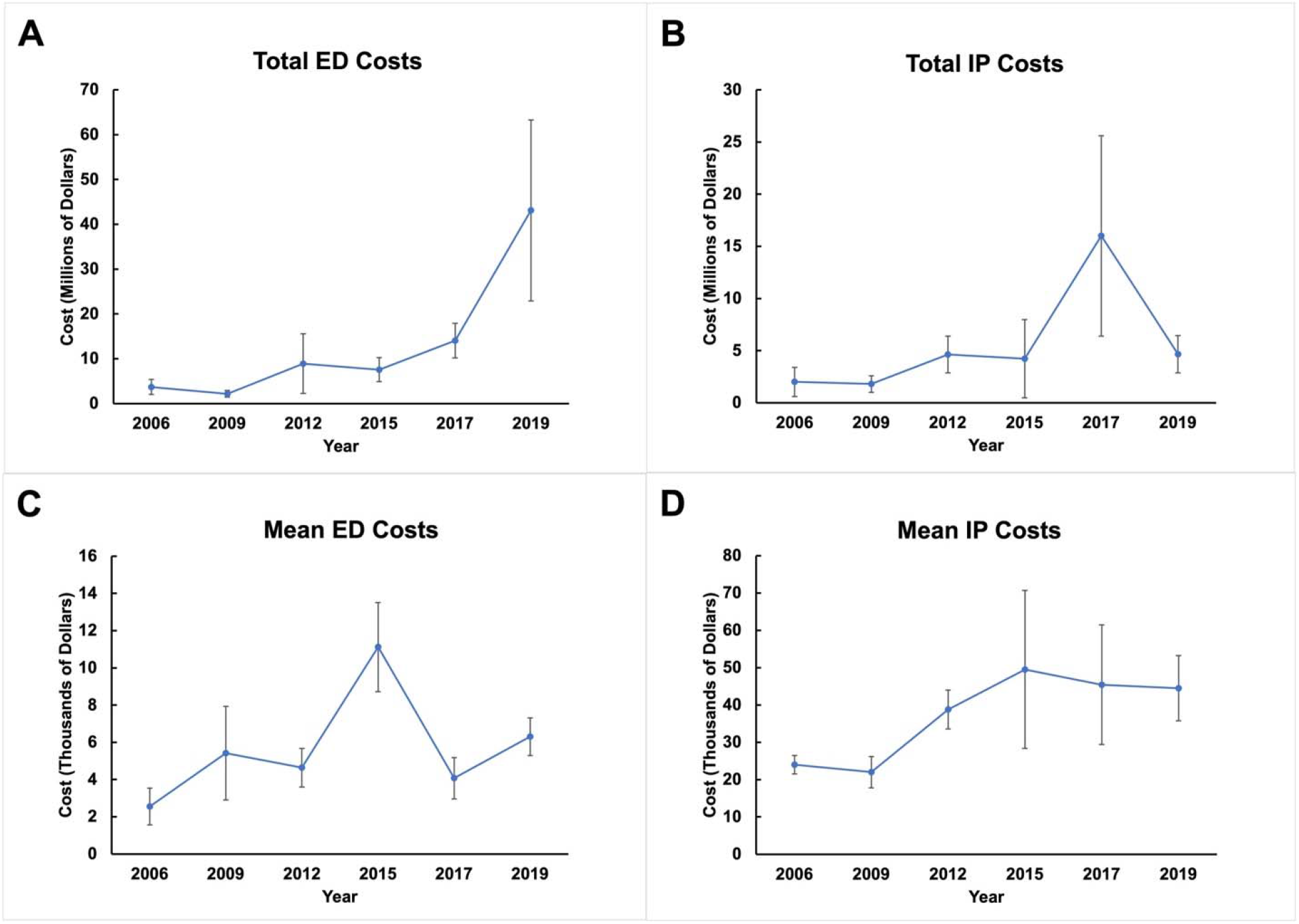
Annual trends of costs for retinal tear patients. A: Total ED costs per year. B: Total IP costs per year. C: Mean ED cost per year. D. Mean IP cost per year. ED: Emergency Department. IP: Inpatient. Error bars represent standard error.

## Discussion

To our knowledge, there is no large-scale study that has studied the epidemiology and economic burden of RTs in the US. Most studies are retrospective chart reviews that characterize the surgical and clinical outcomes of RT.^5-10^ They do not extrapolate to the US population or provide insights into temporal trends of incidence and medical costs. In this study, we identified a stable RT incidence and a decreasing rate of RRDs in primary RT patients. In contrast, ED and IP costs for primary RTs have alarmingly increased during this time period. Altogether, this study raises awareness toward the increasing costs of RTs and the demand to help mitigate these increases.

Within the study time period, primary RT incidence did not significantly change from an annual average of 8.2 per million population. To our knowledge, this is the first report of retinal tear incidence for the US. Compared to RRD studies, our calculated annual RT incidence is smaller than annual RRD incidences, which can range from 91 to 179 per million US population.^11^ Our smaller RT incidence (compared to RRD) indicates that most patients with a RT present to a clinic rather than ED. This makes sense since same-day outpatient clinic use is cheaper than ED use.^17^ Regardless, using ED-level RT encounters is an important way to track treatment costs and outcomes. Most of the RT and RRD studies have been retrospective chart reviews and, hence, do not extrapolate to the entire nation. Furthermore, they do not provide an insight into the treatment cost for these patients.^5-10^

The stable trend of RT incidence reported in our study mirrors the global trend (from 2000 to 2020) of myopia and high myopia incidences reported in a previous study.^18^ In contrast to the time period between 2000 and 2020, Holden and colleagues^18^ estimate that myopia incidence will be significantly higher starting 2030 and increase by 238% by 2050 (compared to 2000). Due to the high number of myopes in the future, the incidence of high-myopia-related problems, such as RTs, RRDs, glaucoma, and lattice degeneration, will greatly increase and present a larger burden to the healthcare system. To ensure that physicians are prepared for the future increase in myopes (and hence RTs and RRDs), ophthalmologists have to become more vigilant to diagnose high-myopia and its downstream issues.

To prepare for the high amount of myopia and myopia-associated issues in the future, we recommend strategies that increase surveillance and access to healthcare. Traditionally, visual acuity exams are common for children in the early education years.^19^ However, in our study, RTs were most commonly found in patients at least 45 years of age (79%). Therefore, we recommend physicians to increase surveillance for RTs and other myopia-related conditions in patients of at least 45 years of age. Furthermore, a significant amount of our RT patient population consists of Hispanics descent, Medicare enrollees, and residents of low-income neighborhoods. It is well known that one’s income, neighborhood, and socioeconomic status are related to one’s access to healthcare and health outcomes.^20^ These demographics are more likely to be uninsured, deal with communication barriers with their physician, and receive lower-quality care.^21–24^ Briefly, to reduce healthcare inequality within the RT population, the physical and financial barrier to an eye professional has to decrease.^24^ Together, these should help RT patients to receive high-quality care.

Interestingly, we found RTs occurred 1.5 times more often in men than women. This relationship has been shown in other studies before. Men are more likely to have retinal breaks after a posterior vitreous detachment, have a higher rate of treatment failure, and have a higher rate of a new RT.^6,9,25^ Along with RTs, men are more likely to have a RRD and require surgery for a RRD.^11,26^ In contrast, research has shown that females are more likely to have a posterior vitreous detachment, which is suspected to be due to a higher amount of advanced glycation end products and steroid differences.^27–29^ It is possible that even though women have more degeneration of the posterior vitreous humor, there is a higher chance of the retina being tugged inward in men. A higher chance of retinal traction could occur if vitreous degeneration in males is closer to the inner surface of the retina, while in females it is closer to the center of the posterior vitreous cavity. Furthermore, it is possible that the collagen fibers in females are more loosely connected to the retina than in males. However, this is very little data on the biochemical difference between male and female vitreous humor.

Despite the stable RT incidence, we have detected a large increase in mean ED and IP charges. This mirrors the increase of healthcare costs found in many different aspects of the medical field. Increases in service price and intensity have contributed 50% and the aging population has contributed 12% to the increase in annual healthcare from 1996 to 2013.^12^ Studies have shown that doctors in high-healthcare-cost regions and low-healthcare-cost regions are equally likely to recommend treatment options that are heavily backed by data. However, doctors in high-healthcare-cost regions are more likely to recommend treatment options that are not as well backed by science and more likely to admit patients into an intensive care unit.^13,30^ Since the Northeast region accounts for almost half of our RT population, it is possible that doctors in these regions are recommending treatment options to RT patients that increase RT treatment intensity, which does not necessarily lead to better outcomes. Furthermore, there has to be changes on the policy level to ensure that care is administered in an efficient way without harming patient outcomes. Research has shown that if care is delivered in an integrated manner (e.g. physicians work in an network and provided team-based care) has great potential of reducing costs.^31^

Although there are many advantages of using NEDS to track RT incidence and economic burden, there are some important limitations to the database and to our study. First, NEDS is an encounter-level database and it does not provide us longitudinal information about a patient’s care. Therefore, if the same RT patient presented to the ED twice in the same year, NEDS would list that as two separate encounters. Furthermore, the database does not include ophthalmology-specific data that is useful in characterizing eye diseases, such as intraocular pressure, visual acuity, location of tears, and size of tears.

In conclusion, despite the stable incidence of RTs, there has been large increases in ED and IP charges. This parallels what has been seen throughout the entire medical field.^12,14,16^ Therefore, we recommend doctors and lawmakers to work together to mitigate these increasing costs.

## Data Availability

The data can be bought from the following link:https://www.hcup-us.ahrq.gov/nedsoverview.jsp

## Acknowledgements

The authors have no relevant acknowledgements to make.

## Figure Captions

**Supplementary Table 1:** Demographics of Patients with a Primary Retinal Tear

| Characteristics | Young Children (0-10) N (%) | Adolescents (11-20) N (%) | Young Adults (21-44) N (%) | Adults (45-65) N (%) | Elderly (≥65) N (%) | Total N (%) |
| --- | --- | --- | --- | --- | --- | --- |
| **Sex** |  |  |  |  |  |  |
| Male | 52 (57.6) | 224 (51.3) | 1415 (59.3) | 5784 (61.3) | 2195 (62.8) | 9670 (61) |
| Female | 39 (42.4) | 213 (48.7) | 973 (40.7) | 3650 (38.7) | 1297 (37.2) | 6171 (39) |
| **Race** |  |  |  |  |  |  |
| White | * | 40 (23.5) | 287 (34.7) | 2048 (50.2) | 1002 (62.4) | 3376 (50.3) |
| Black | * | 49 (28.9) | 198 (24) | 352 (8.6) | 122 (7.6) | 722 (10.8) |
| Hispanic | 21 (82.4) | 72 (41.8) | 292 (35.4) | 1517 (37.2) | 444 (27.7) | 2346 (35) |
| Asian or Pacific Islander | * | * | 20 (2.4) | 63 (1.6) | 17 (1.1) | 106 (1.6) |
| Native American | * | * | * | 11 (0.3) | * | 11 (0.2) |
| Other | * | * | 29 (3.6) | 89 (2.2) | 19 (1.2) | 147 (2.2) |
| **Primary Payment Method** |  |  |  |  |  |  |
| Medicare | * | * | 43 (1.8) | 519 (5.5) | 2459 (71.2) | 3021 (19.2) |
| Medicaid | 56 (65.8) | 163 (37.4) | 472 (20) | 775 (8.3) | 117 (3.4) | 1582 (10.1) |
| Private Insurance | 13 (15.5) | 161 (37) | 1090 (46.2) | 5797 (61.8) | 688 (19.9) | 7750 (49.3) |
| Self-Pay | * | 86 (19.7) | 579 (24.5) | 1622 (17.3) | 122 (3.5) | 2419 (15.4) |
| No Charge |  |  | * | 23 (0.2) |  | 33 (0.2) |
| Other | * | 26 (6) | 165 (7) | 648 (6.9) | 69 (2) | 915 (5.8) |
| **Median Household Income in Patient's Zipcode** |  |  |  |  |  |  |
| 1<sup>st</sup> Quartile | 27 (31.6) | 115 (27.8) | 652 (28.4) | 1896 (21.3) | 741 (22.2) | 3430 (22.8) |
| 2<sup>nd</sup> Quartile | 11 (13.1) | 96 (23) | 537 (23.4) | 2037 (22.9) | 851 (25.5) | 3532 (23.5) |
| 3<sup>rd</sup> Quartile | 19 (22) | 65 (15.8) | 610 (26.5) | 2303 (25.8) | 833 (24.9) | 3829 (25.4) |
| 4<sup>th</sup> Quartile | 28 (33.4) | 139 (33.4) | 500 (21.8) | 2673 (30) | 919 (27.5) | 4260 (28.3) |
| **Associated Comorbidities** |  |  |  |  |  |  |
| Hyperlipidemia | * | * | 23 (1) | 455 (4.8) | 366 (10.5) | 844 (5.3) |
| Diabetes | * | * | 75 (3.1) | 670 (7.1) | 395 (11.3) | 1139 (7.2) |
| Overweight | * | * | 38 (1.6) | 142 (1.5) | 46 (1.3) | 227 (1.4) |
| Hypertension | * | * | 136 (5.7) | 1774 (18.8) | 1094 (31.3) | 3004 (19) |
| Degenerative | * | 22 (4.9) | 69 (2.9) | 103 (1.1) | 18 (0.5) | 211 (1.3) |
| **Myopia** |  |  |  |  |  |  |
| **Myopia** | * | 31 (7.2) | 99 (4.2) | 233 (2.5) | 33 (0.9) | 402 (2.5) |
| **Cataract** | * | 15 (3.5) | 152 (6.4) | 1609 (17.1) | 570 (16.3) | 2352 (14.8) |
| **Glaucoma** | * | * | 115 (4.8) | 211 (2.2) | 178 (5.1) | 508 (3.2) |
| **Tobacco** | * | * | 80 (3.4) | 597 (6.3) | 414 (11.8) | 1091 (6.9) |
| **Alcohol** | * | * | 13 (0.5) | 24 (0.3) | * | 41 (0.3) |
| **Rhegmatogenous Retinal Detachment** | 68 (74.3) | 309 (70.9) | 1796 (75.2) | 5947 (63) | 2457 (70.4) | 10577 (66.8) |
| **ED Outcome** |  |  |  |  |  |  |
| **Admitted Inpatient** | * | 30 (6.9) | 173 (7.3) | 491 (5.2) | 126 (3.6) | 826 (5.2) |

## References

1. Yonemoto J, Noda Y, Masuhara N, Ohno S. Age of onset of posterior vitreous detachment. Curr Op in Ophthalmol. 1996;7(3):73–76. doi:10.1097/00055735-199606000-00012

2. Foos RY, Wheeler NC. Vitreoretinal juncture. Synchysis senilis and posterior vitreous detachment. Ophthalmol. 1982;89(12):1502–1512. doi:10.1016/s0161-6420(82)34610-2

3. Morita H, Funata M, Tokoro T. A CLINICAL STUDY OF THE DEVELOPMENT OF POSTERIOR VITREOUS DETACHMENT IN HIGH MYOPIA. Retina. 1995;15(2):117–124. doi:10.1097/00006982-199515020-00005

4. Hikichi T, Akiba J, Trempe CL. Prevalence of posterior vitreous detachment in retinitis pigmentosa. Ophthalmic Surg. 1995;26(1):34–38. https://www.ncbi.nlm.nih.gov/pubmed/7746622

5. Byer NE. Natural history of posterior vitreous detachment with early management as the premier line of defense against retinal detachment. Ophthalmol. 1994;101(9):1503–1513; discussion 1513-1514. doi:10.1016/s0161-6420(94)31141-9

6. Novak MA, Welch RB. Complications of acute symptomatic posterior vitreous detachment. Am J Ophthalmol. 1984;97(3):308–314. doi:10.1016/0002-9394(84)90628-7

7. Shunmugam M, Ang GS, Lois N. Giant retinal tears. Surv Ophthalmol. 2014;59(2):192–216. doi:10.1016/j.survophthal.2013.03.006

8. Sharma MC, Regillo CD, Shuler MF, Borrillo JL, Benson WE. Determination of the incidence and clinical characteristics of subsequent retinal tears following treatment of the acute posterior vitreous detachment-related initial retinal tears. Am J Ophthalmol. 2004;138(2):280–284. doi:10.1016/j.ajo.2004.03.009

9. Brod RD, Lightman DA, Packer AJ, Saras HP. Correlation between vitreous pigment granules and retinal breaks in eyes with acute posterior vitreous detachment. Ophthalmol. 1991;98(9):1366–1369. doi:10.1016/s0161-6420(91)32124-9

10. Sarrafizadeh R, Hassan TS, Ruby AJ, et al. Incidence of retinal detachment and visual outcome in eyes presenting with posterior vitreous separation and dense fundus-obscuring vitreous hemorrhage. Ophthalmol. 2001;108(12):2273–2278. doi:10.1016/s0161-6420(01)00822-3

11. Mitry D, Charteris DG, Fleck BW, Campbell H, Singh J. The epidemiology of rhegmatogenous retinal detachment: geographical variation and clinical associations. Br J Ophthalmol. 2010;94(6):678–684. doi:10.1136/bjo.2009.157727

12. Dieleman JL, Squires E, Bui AL, et al. Factors Associated With Increases in US Health Care Spending, 1996-2013. JAMA. 2017;318(17):1668–1678. doi:10.1001/jama.2017.15927

13. Fisher ES, Bynum JP, Skinner JS. Slowing the growth of health care costs--lessons from regional variation. N Engl J Med. 2009;360(9):849–852. doi:10.1056/NEJMp0809794

14. Lane BH, Mallow PJ, Hooker MB, Hooker E. Trends in United States emergency department visits and associated charges from 2010 to 2016. Am J Emerg Med. 2020;38(8):1576–1581. doi:10.1016/j.ajem.2019.158423

15. Iftikhar M, Latif A, Usmani B, Canner JK, Shah SMA. Trends and Disparities in Inpatient Costs for Eye Trauma in the United States (2001-2014). Am J Ophthalmol. 2019;207:1–9. doi:10.1016/j.ajo.2019.05.021

16. Iftikhar M, Canner JK, Hall L, Ahmad M, Srikumaran D, Woreta FA. Characteristics of Orbital Floor Fractures in the United States from 2006 to 2017. Ophthalmol. 2021;128(3):463–470. doi:10.1016/j.ophtha.2020.06.065

17. Singman EL, Smith K, Mehta R, et al. Cost and Visit Duration of Same-Day Access at an Academic Ophthalmology Department vs Emergency Department. JAMA Ophthalmol. 2019;137(7):729. doi:10.1001/jamaophthalmol.2019.0864

18. Holden BA, Fricke TR, Wilson DA, et al. Global Prevalence of Myopia and High Myopia and Temporal Trends from 2000 through 2050. Ophthalmol. 2016;123(5):1036–1042. doi:10.1016/j.ophtha.2016.01.006

19. Anstice NS, Thompson B. The measurement of visual acuity in children: an evidence□based update. Clin and Exp Opto. 2014;97(1):3–11. doi:10.1111/cxo.12086

20. Lurie N, Dubowitz T. Health disparities and access to health. JAMA. 2007;297(10):1118–1121. doi:10.1001/jama.297.10.1118

21. Artiga S, Hill L, Orgera K, Damico A. Health coverage by race and ethnicity, 2010-2019. KFF. Published July 16, 2021. Accessed July 26, 2022. https://www.kff.org/racial-equity-and-health-policy/issue-brief/health-coverage-by-race-and-ethnicity/

22. Aboul-Enein FH, Ahmed F. How language barriers impact patient care: a commentary. J Cult Divers. 2006;13(3):168–169. https://www.ncbi.nlm.nih.gov/pubmed/16989255

23. Al Shamsi H, Almutairi AG, Al Mashrafi S, Al Kalbani T. Implications of Language Barriers for Healthcare: A Systematic Review. Oman Med J. 2020;35(2):e122. doi:10.5001/omj.2020.40

24. Eisenberg JM, Power EJ. Transforming insurance coverage into quality health care: voltage drops from potential to delivered quality. JAMA. 2000;284(16):2100–2107. doi:10.1001/jama.284.16.2100

25. Smiddy WE, Flynn HW, Nicholson DH, et al. Results and Complications in Treated Retinal Breaks. Am J of Ophthalmol. 1991;112(6):623–631. doi:10.1016/s0002-9394(14)77267-8

26. Wong TY, Tielsch JM, Schein OD. Racial difference in the incidence of retinal detachment in Singapore. Arch Ophthalmol. 1999;117(3):379–383. doi:10.1001/archopht.117.3.379

27. Hayashi K, Sato T, Manabe SI, Hirata A. Sex-Related Differences in the Progression of Posterior Vitreous Detachment with Age. Ophthalmol Retina. 2019;3(3):237–243. doi:10.1016/j.oret.2018.10.017

28. van Deemter M, Ponsioen TL, Bank RA, et al. Pentosidine accumulates in the aging vitreous body: A gender effect. Exp Eye Res 2009;88(6):1043–1050. doi:10.1016/j.exer.2009.01.004

29. Nishikawa Y, Morishita S, Horie T, et al. A comparison of sex steroid concentration levels in the vitreous and serum of patients with vitreoretinal diseases. PLoS One. 2017;12(7):e0180933. doi:10.1371/journal.pone.0180933

30. Sirovich B, Gallagher PM, Wennberg DE, Fisher ES. Discretionary decision making by primary care physicians and the cost of U.S. Health care. Health Aff. 2008;27(3):813–823. doi:10.1377/hlthaff.27.3.813

31. Rocks S, Berntson D, Gil-Salmerón A, et al. Cost and effects of integrated care: a systematic literature review and meta-analysis. Eur J Health Econ. 2020;21(8):1211–1221. doi:10.1007/s10198-020-01217-5

